# Importance of external validation and subgroup analysis in artificial-intelligence detecting low ejection fraction from electrocardiograms

**DOI:** 10.1101/2022.07.22.22277758

**Authors:** Ryuichiro Yagi, Shinichi Goto, Yoshinori Katsumata, Calum A MacRae, Rahul C Deo

## Abstract

**Introduction:** Asymptomatic left ventricular dysfunction (ALVD) carries an increased risk for overt heart failure and mortality, yet treatable to mitigate disease progression. An artificial intelligence (AI)-enabled 12-lead electrocardiogram (ECG) model demonstrated promise in ALVD screening but an unexpected drop in performance was observed in external validation. We thus sought to train a *de novo* model for ALVD detection and investigate its performance across multiple institutions and across a broader set of patient strata.

**Methods:** ECGs taken within 14 days of an echocardiogram were obtained from 4 academic hospitals from the United States (BWH, MGH and UCSF) and Japan (Keio). Four AI models were trained to detect patients with ejection fraction < 40% using ECG from each of the 4 institutions. All the models were then evaluated on the held-out test dataset from the same intuition and all data from the 3 external institutions. Subgroup analyses stratified by patient characteristics and common overt ECG abnormalities were performed.

**Results:** A total of 221,846 ECGs were identified from the 4 institutions. While the BWH-trained and Keio-trained model yielded similar accuracy on its internal test data (AUROC 0.913 and 0.914 respectively), external validity was worse for Keio-trained model (AUROC: 0.905-0.915 for BWH-trained and 0.849-0.877 for Keio-trained model). Although ECG abnormalities including atrial fibrillation, left bundle branch block and paced rhythm reduced detection, the models performed robustly across patient characteristics and other ECG features.

**Conclusion:** Different dataset produces models with different performance highlighting the importance of external validation and extensive stratification analysis.

## Introduction

Asymptomatic left ventricular dysfunction (ALVD) is a frequently observed finding that carries an increased risk for overt heart failure and mortality(1). If detected early, ALVD can be treated to mitigate disease progression. An artificial intelligence (AI)-enabled 12-lead electrocardiogram (ECG) model has demonstrated promise in ALVD screening(2) but an unexpected drop in performance was observed in external validation(3). In addition to problems with generalizability, AI models have also shown uneven performance in distinct subpopulations(4), which has important implications for downstream decision-making should these models be applied to general practice. We thus sought to train a *de novo* model for ALVD detection from ECG data and investigate its performance across multiple institutions and across a broader set of patient strata.

## Methods

ECGs taken within 14 days of an echocardiogram in patients aged ≥ 20 were obtained from 4 academic hospitals from the United States (Brigham and Women’s Hospital (BWH), Massachusetts General Hospital (MGH), University of California San Francisco (UCSF)) and Japan (Keio University Hospital). A convolutional neural network (CNN) was trained to detect patients with left ventricular ejection fraction < 40% from ECG alone. The dataset for each institution was randomly divided into three groups (derivation, validation, and test) in a 5:2:3 ratio without overlaps of patients across groups. Models were trained on the derivation dataset and those with highest performance on the validation dataset across 50 epochs were chosen as the final model. These models were then evaluated on the test dataset from the same intuition and all data from the 3 external institutions. While training was performed using all ECGs, the models were tested using a single ECG-echocardiogram pair with the closest dates for each patient. Subgroup analyses stratified by patient characteristics and common overt ECG abnormalities were performed. The model performance was evaluated by area under the receiver operating characteristics curve (AUROC) analysis.

## Results

There were 75,033, 79,663, 36,314 and 30,836 ECGs for BWH, MGH, UCSF and Keio, respectively. While the BWH-trained model yielded excellent accuracy on internal test data (AUROC 0.913 [95%CI, 0.901-0.926]) and good external validity (AUROC 0.905 [0.880-0.905], 0.910 [0.893-0.928], and 0.915 [0.895-0.935]), respectively for MGH, UCSF and Keio), the Keio-trained model, having a similar performance on its test set (AUROC 0.914 [0.893-0.936]), showed poor external validity (AUROC 0.856 [0.838-0.875], 0.849 [0.826-0.872], and 0.877 [0.856-0.897], respectively for BWH, MGH and UCSF) (**Figure 1A**). The stratified analysis, using the BWH-trained model, demonstrated consistent performance across patient age, sex, race and common ECG abnormalities: first degree atrio-ventricular block and right bundle brunch block (AUROC 0.960 [0.928-0.992], and 0.883 [0.819-0.948], respectively). However, the model showed lower accuracy in individuals with atrial fibrillation (AF), left bundle branch block (LBBB), or a paced rhythm (AUROC 0.856 [0.821-0.892], 0.791 [0.717-0.866], and 0.859 [0.818-0.899], respectively) (**Figure 1B**). These findings were consistent upon external validation.

**Figure 1.**
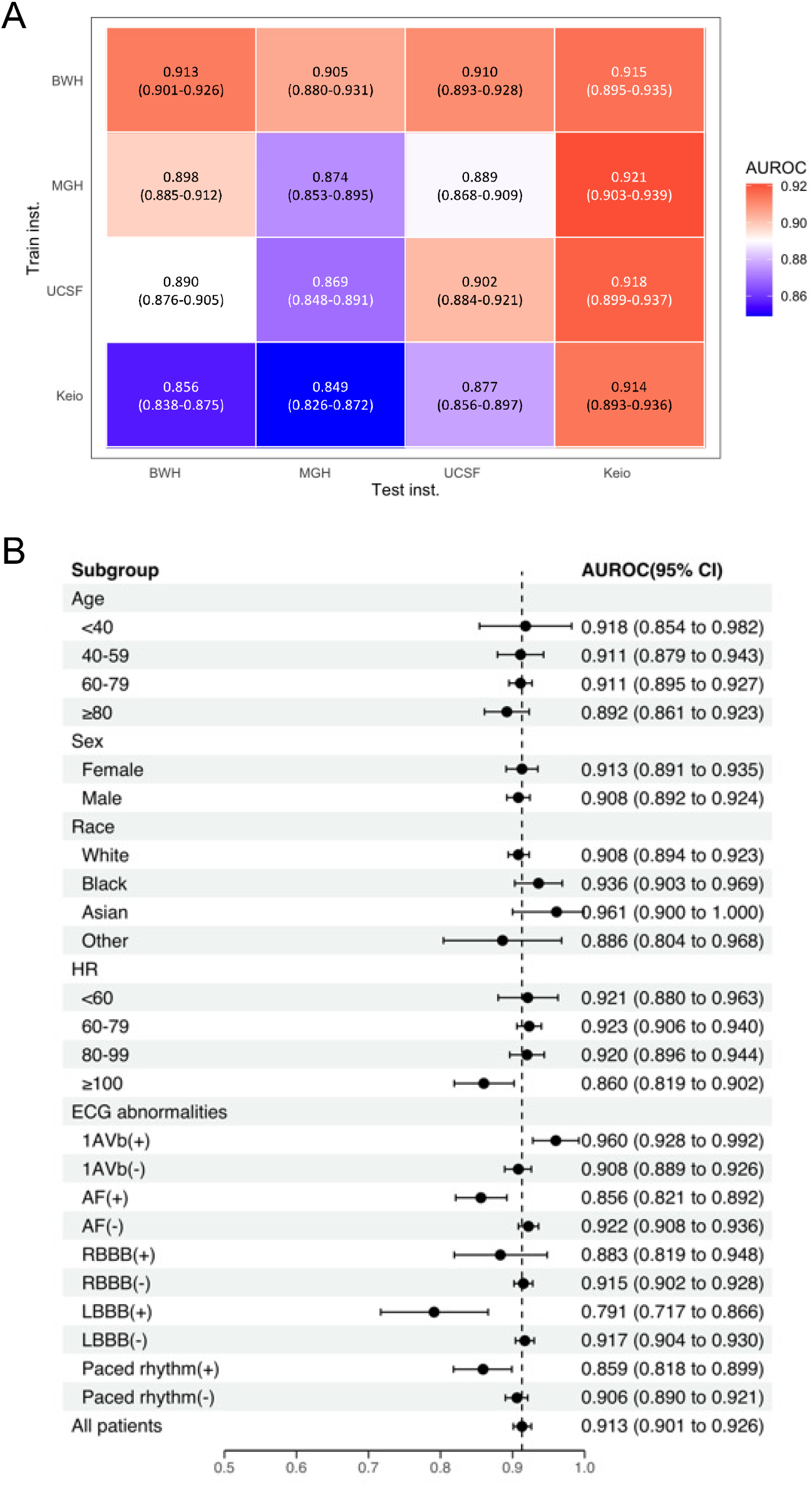
**A**. Performance and external validity of models trained at individual institution. **B**. Stratification analyses on patient characteristics and ECG abnormalities in BWH. HR: heart rate, AF: atrial fibrillation, CLBBB: complete left bundle branch block, PM: pacemaker.

## Discussion

Although a previously published model reported high performance on an internal dataset, retrospectively and prospectively(2, 5), external validation revealed an unexpectedly poor performance(3). In our analyses, the models trained at BWH and Keio both displayed similar performance on internal test sets but varied substantially upon external validation. If only internal testing had been performed, one could have concluded these models were equally useful. Furthermore, although published ALVD detection models appear robust across patient demographics(2, 6), our subgroup analysis by ECG abnormalities demonstrated settings in which the model had diminished performance, which was confirmed upon external validation. Given that patients with newly-detected AF and LBBB are likely to be referred for echocardiography, regardless, the models’ high accuracy for other subgroups imply excellent utility for screening. These findings highlight the importance of extensive stratification analysis and external validation to establish model applicability.

## Data Availability

The code for training and testing the model is provided at https://github.com/obi-ml-public/ECG-LV-Dysfunction. The model weights may contain personal information from patients and thus, are not shared. We provide a web-interface to run our model and generate predictions at http://onebraveideaml.org

## Acknowledgements

This work was supported by One Brave Idea, co-funded by the American Heart Association and Verily with significant support from AstraZeneca and pillar support from Quest Diagnostics. R.C.D is supported by grants from the National Institute of Health, the American Heart Association (One Brave Idea, Apple Heart and Movement Study), has received consulting fees from Novartis and Pfizer, and is co-founder of Atman Health. C.A.M. is a consultant for Pfizer and co-founder of Atman Health. SG is partially supported by Drs. Morton and Toby Mower Science Innovation Fund Fellowship, a grant from The Japanese Society of Thrombosis and Hemostasis and One Brave Idea. This study complies with all ethical regulations and guidelines. Approval was obtained from the Institutional Review Boards of all institutions.

